# A Multilevel Intervention to Reduce Chronic Disease Risk in Socioeconomically Disadvantaged Unemployed Populations: The *North Carolina Works 4 Health (NCW4H)* Clinical Trial Protocol

**DOI:** 10.1101/2024.01.11.24301167

**Authors:** Shawn M. Kneipp

**Author notes:** Corresponding Author: Shawn Kneipp UNC-Chapel Hill School of Nursing Carrington Hall, CB# 7460 Chapel Hill, NC 27599.

## Abstract

**Background:** The overall goal of the North Carolina Works for Health (NCW4H) study is to adapt and test the effectiveness of a multilevel intervention to reduce chronic disease risk in socioeconomically disadvantaged, unemployed (SEDU) populations who rely on publicly-funded job placement programs to secure employment. Studies have shown an unemployment episode increases in psychological distress, health-compromising coping behavior, blood pressure, and weight gain – all of which increase chronic disease risk.

**Methods:** A randomized, 2 x 2 factorial design will test an individual level (IL) and employer level (EL) intervention, and their joint effects, in SEDU adults receiving job placement services through publicly-funded programs. Interventions consist of a chronic disease prevention program adapted from the Diabetes Prevention Program at the IL, and an implicit bias-based supervisor support program for newly-hired SEDU adults at the EL. We will enroll 600 SEDU adults 18 to 64 years of age who have either received public assistance benefits in the prior two years *or* have <4-year college degree, used publicly-funded job placement services during the most recent unemployment episode, and are not receiving disability income, not pregnant, and fluent in English; and 80-200 supervisors of our SEDU enrolled participants when hired by an employer. Primary outcomes include psychological distress, blood pressure, and weight gain, and will be collected at baseline and 3, 6, and 12 months post-enrollment. Secondary outcomes related to coping, health behavior, workplace support, and employment will also be collected. Main effects of the IL and EL interventions, and IL x EL interactions, will be analyzed using generalized multivariate models accounting for clustering effects.

**Discussion:** This multilevel intervention is novel in that it is designed to mitigate chronic disease risk during an unemployment episode, and includes an intervention at a level at the employer level, where social determinants of health operate. Despite the design challenges that multilevel intervention trials such as the NCW4H study present, they are needed to meaningfully address health inequities in the U.S. Findings from this study are expected to inform how approaches that incorporate public health and employment sectors could reduce chronic disease among socioeconomically disadvantaged populations.

**Ethics and dissemination:** The trial is ongoing and has been approved by the University of North Carolina at Chapel Hill Institutional Review Board. Trial results will be published in a scientific journal and presented at scientific conferences.

**Trial registration number:** NCT04815278, ClinicalTrials.gov

## Background and rationale {6a}

Recognized as a key SDOH, employment is a primary source for both economic and social benefits [1]. A single unemployment episode precipitates a cascade of adverse psychological, emotional, coping, and behavioral sequelae [2–4]. A high rate of psychological distress follows an unemployment episode [2, 5, 6], where there is a two- to three-fold increase in the risk of developing depression [4, 6]. The stress from unemployment can also precipitate a reliance on avoidance- and emotion-focused coping strategies that result in less physical activity, consuming a less healthy diet, and increased substance use [3, 7, 8]. In turn, these behavioral risks following an unemployment episode accelerate weight gain (averaging 15-20 pounds) [9], increase the odds of developing a chronic disease by 43% [10], and increase the risk of death by 73-77% [11]. Notably, these unemployment-related health risks persist for up to 10 years post-job loss – regardless of the length of the unemployment episode, regaining employment, or the overall strength of the economy [12]. The COVID-19 pandemic extended the reach of unemployment and its consequences to groups with previously stable employment, where the unemployment rate peaked at 14.8% in April 2020, surpassing the rate of 10% during the 2007 Great Recession [13].

The impacts of unemployment and chronic disease inequities are felt acutely in the everyday lives of socioeconomically disadvantaged (SED) populations (i.e., people with lower income, lower education, and/or who are racial/ethnic minorities). Only 30% of lower-wage workers in the U.S. are eligible for unemployment benefits, and other economic safety net programs are becoming less secure, requiring job search engagement or some form of work for benefit receipt, largely through Department of Social Services Employment and related publicly-funded programs (DSS-E) [14]. Working age people who are SED are more than twice as likely to be unemployed than their more advantaged counterparts [15, 16] – a gap that was exacerbated during the COVID-19 pandemic [13]. Inequities in the burden of chronic disease prevalence, morbidity, and mortality are also higher among SED groups in the U.S. than in the general population [17], which can result from disadvantage in the labor market and hamper securing or maintaining employment.

Chronic disease risk accrues not only based on employment or unemployment status, but also from physical and psychosocial factors in the workplace [18]. Psychosocial factors at work such as job control, effort-reward imbalance, and supervisor support are significant predictors of a wide range of health outcomes [19–21]. The lower-wage workforce is largely composed of individuals who are SED, precariously employed, and report less job control and supervisor or other support in the workplace than higher-income earners [22, 23]. With increased attention to employment as a SDOH over the past decade, the critical role of supervisors in promoting the psychosocial health of employees has gained attention [24]. Supervisor support is negatively associated with employee stress [25], presenteeism [26], and turnover intention [27]; positively associated with mental health [27, 28], work engagement [29], adhering to safety practices [30]; and can mediate the effect of job uncertainty on emotional exhaustion [31]. Interventions to build family-supportive supervisor behaviors have been shown to improve employee blood pressure [32], job satisfaction [27], and reduce psychological distress and burnout [27].

Supervisors, however, also bring implicit (or unintentional) bias to their roles at work [33]. Negative bias toward SED workers, including those receiving job search or placement support through DSS-E programs (sometimes referred to as ‘welfare bias’) [34] can shape supervisors’ expectancies of poor performance, that, ironically, can contribute to actual poor work performance and make succeeding in a new job difficult for SED workers hired following an unemployment episode [35].

Taken together, the stress-chronic disease cascade initiated during an unemployment episode can be exacerbated by moving into a job with a less supportive environment. For newly hired individuals from SED backgrounds who have received job search assistance through DSS-E programs, having less supervisor support due to a supervisor’s expectancy bias of poor performance can result in recurring unemployment.

Although several studies have tested health promotion programs for lower-wage workers in the workplace [36], fewer have tested interventions to improve the health of unemployed populations [37]. Moreover, with the exception of supported employment programs designed for job seekers with a disabling mental illness [38], intervention research in this area has not focused on the unemployment-to-employment transition experience, where interventions have been designed to interact across two or more levels (individual, organizational, etc.,) as SED individuals who are at high risk for chronic disease morbidity and mortality transition from unemployment into employment. To adequately address the complexity of the individual and joint contributions of interventions across levels to address SDOH, novel prevention-focused, public health approaches [39] that include cross-sector partnerships are needed.

## Objectives {7}

The overall goal of the *North Carolina Works 4 Health* (NCW4H) study is to adapt (Phase I) and test (Phase II) the effectiveness of a multilevel intervention (MLI) that targets psychological and behavioral factors at the individual-level (IL) through the *Chronic Disease Prevention Program* (CDPP), and supervisor support at the employer-level (EL) through *the Supervising for Success Program* (S4S) (see Figure 1).

**Figure 1.**
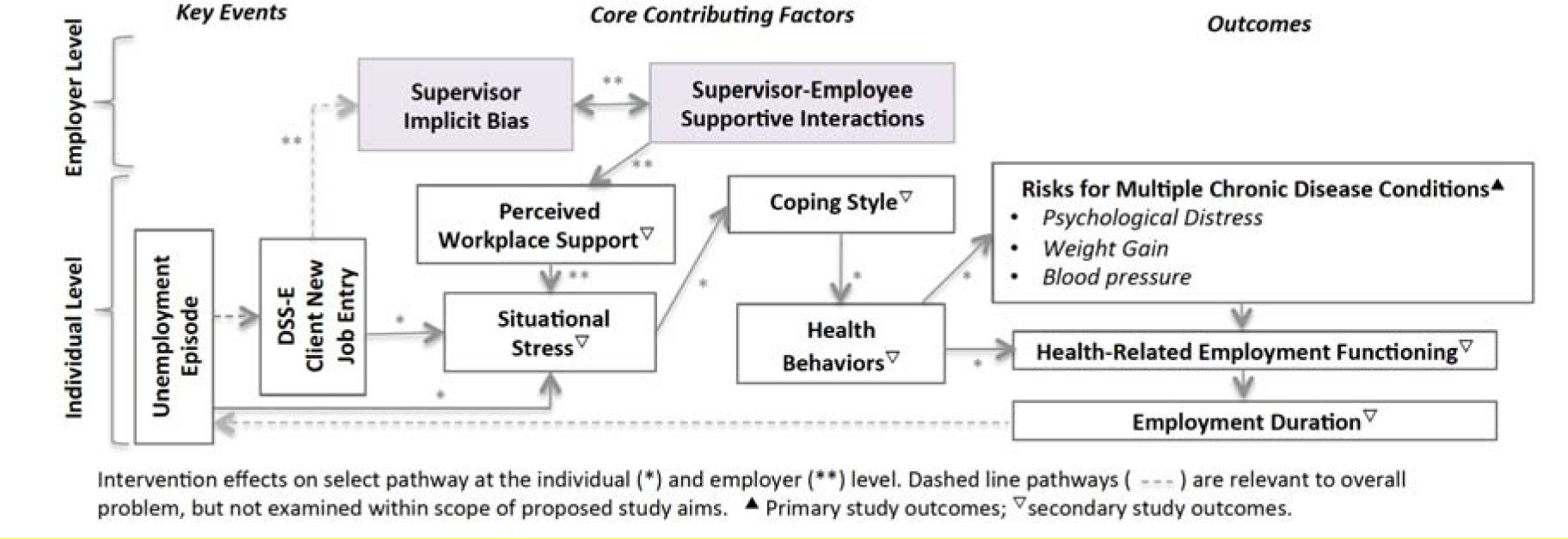
Chronic Disease Risk and Intervention Effect Pathways on Primary and Secondary Outcomes of the NCW4H Trial.

## Trial Design {8}

The RCT portion (Phase II) of the NCW4H study uses a 2×2 factorial design (Figure 2), while also accounting for the naturally-occuring trajectory of an unemployment episode and the chronic disease risk accrual that is initiated and potentially exacerbated after moving into a new job setting. Unemployed, SED individuals are randomized at the IL to the CDPP or a control group using a permuted block approach (in blocks of 4), and monitored monthly for when they begin employment. When employment is found by participants enrolled at the IL, the initial protocol was to first enroll and randomize their employer using a biased coin approach, and then assign the supervisor (nested within employer) to either receive the *S4S* intervention or control condition (no intervention). Intervention delivery of the S4S program to supervisors is time sensitive (to occur shortly after IL participant is hired) and there were unanticipated delays in contacting employers in a post-COVID labor market [40]. Given this, the EL design was modified early in the trial based on employer feedback that all employers desired access to the intervention, and changes in study recruitment materials for employers emphasizing the intervention’s potential to improve retention and meet diversity, equity, and inclusion professional development goals for supervisors.

**Figure 2.**
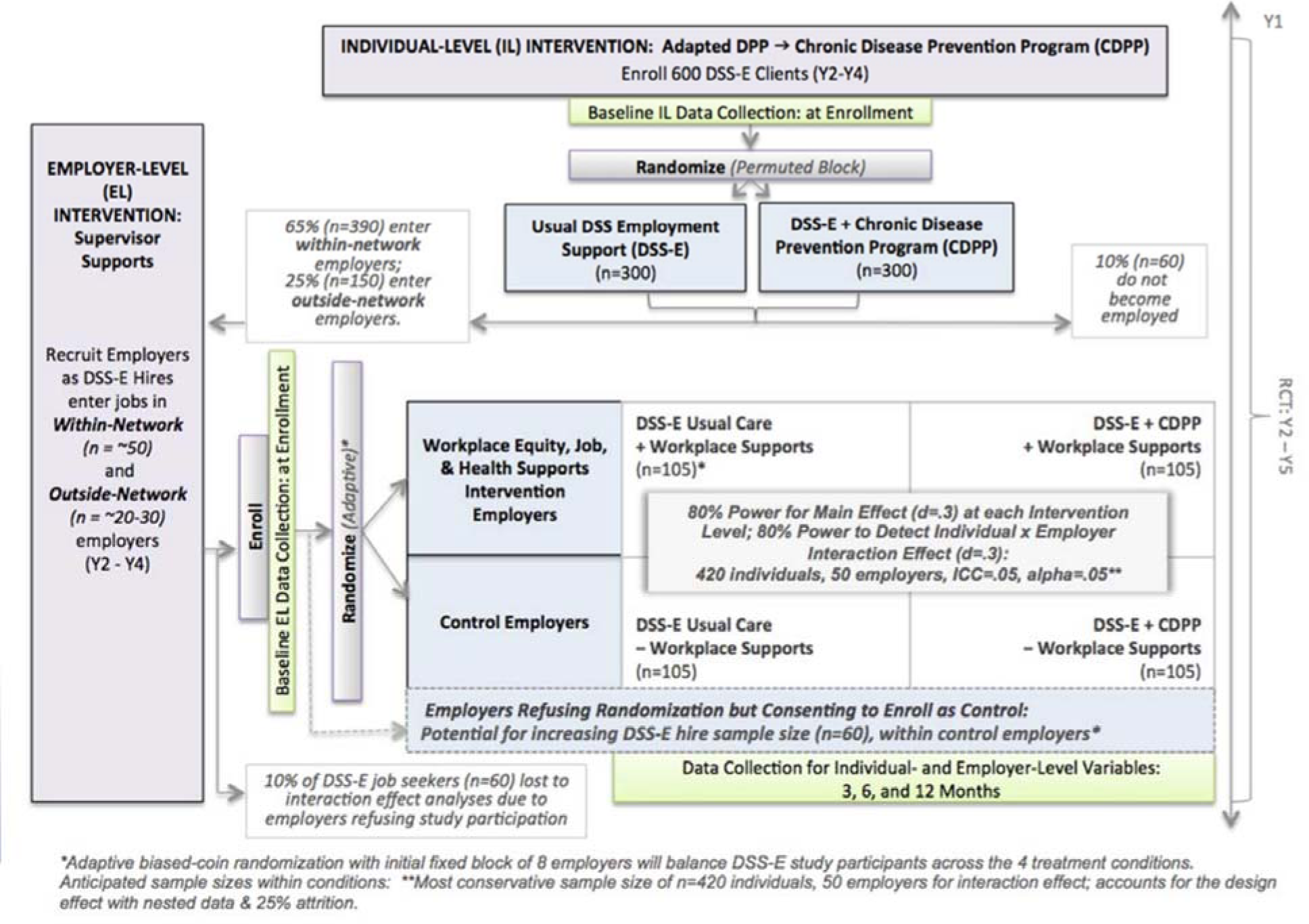
Multilevel NC Works4Health Intervention Study Design.

The revised protocol includes two strategies: 1) recruit all employers in study counties prior to the employer hiring an IL study participant, and randomize them to receive the intervention immediately, in 3 months, or in 6 months using a stepped wedge approach with the expectation that a moderate number of IL study participants would be hired into these organizations based on prior experience of workforce partners; and 2) recruit and enroll individual supervisors when identified by newly hired IL study participants, with those not already embedded in an employer organization being randomized using a permuted block approach (in blocks of 4).

## Methods: participants, interventions, and outcomes

### Study setting {9}

This is a community-based trial taking place in 8 adjacent counties in central North Carolina (Orange, Wake, Chatham, Alamance, Guilford, Durham, Caswell, and Person counties). Study counties comprise both rural and urban areas.

### Eligibility criteria {10}

At the individual-level (IL), eligible participants must be unemployed and looking for work; between the age of 18 and 64 years old; fluent in able to read English; have less than a 4-year college degree or have received any form of public assistance in the prior two years (this includes rental/housing assistance, heat or utility assistance, Supplemental Nutrition Assistance Program (SNAP) benefits, Temporary Assistance for Needy Families (TANF) benefits, or accessing a food pantry); and accessing any resources in study counties available through Department of Social Services Employment and related publicly-funded programs, such as NC Works Career Centers or other publicly-funded job fairs (henceforward, all referred to as DSS-E services) to facilitate job search or job placement activities in the past year. Exclusion criteria include receiving or having a disability benefit case pending; pregnancy; a gastrointestinal condition that requires following strict dietary guidelines; another health condition that significantly limits routine physical activities (e.g., walking); and having any of the following chronic health conditions: severe hypertension with a reading of 180/110 or higher in the past 6 months, a health condition or injury that affects balance or gait, a history of falling in the past 6 months, cancer that is actively being treated with chemotherapy or radiation to the chest or abdomen, inflammatory bowel disease, or and implanted cardiac defibrillator. Inclusion criteria for employers include having a primary or affiliate business in one of the study counties. Individually-enrolled supervisors must be identified as being the supervisor of record for newly-hired, IL study participants.

Given the physical changes that occur with pregnancy related to weight gain, mood, caloric intake, and (potentially) activity, IL participants who become pregnant during the study are excluded from further participation in interventions or data collection, and have the option of either restarting the study from baseline (remaining in their previously allocated group) 6 weeks post-delivery or the end of the pregnancy, or dropping from the study but receiving access to the CDPP intervention modules (as do participants in the IL control group after their 12-month data collection).

### Who will take informed consent? {26a}

Consents at each level, for each recruited group are embedded within an online Qualtrics survey platform. At the IL, a 3-minute video is included as an adjunct option for viewing that explains key aspects of the study using simple terms and graphics designed to improve comprehension. Following review of the online consent, individuals are given contact information if there are any questions prior to selecting their desire to proceed with enrollment, or not. The EL consent is completed online by an administrative official with signing authority within the employing organization, emphasizing the study team will not provide information back on supervisor S4S completion information to the employer. Supervisors identified as supervising newly-hired IL study participants also complete an online consent for enrollment. All consents include contact information for study personnel trained in obtaining consent from human subjects through Institutional Review Board requirements.

### Additional consent provisions for collection and use of participant data and biological specimens {26b}

This trial does not include collecting biological specimens.

## Interventions

### Explanation for the choice of comparators {6b}

Comparators for each main effect of interventions are usual care. At the IL, usual care refers to the job search assistance provided through DSS-E services at the county level, which are similar across study counties. Typically, there are no chronic disease prevention services provided in these settings, nor are job seekers referred to health-related resources explicitly intended to reduce stress or other factors that can increase chronic disease risk. At the EL, usual care refers to professional development and training provided to supervisors within the employer organization, and the professional development and training that individual supervisors may have received within their current or prior employment setting. Exposure to professional development and training across employer organizations and supervisors varies widely; however, as described in the next section, the S4S program was designed to sensitize supervisors to the unique challenges faced by new hires who are SED, and the ways in which implicit bias, often in the form of ‘welfare bias’ (given the reliance of the unemployed SED population on publicly-funded programs to find employment), can surface during supervisor-employee interactions.

## Intervention description {11a}

### Intervention development

#### Individual-Level Intervention: CDPP

Phase I of the *NC Works 4 Health* study included adapting existing, evidence-based interventions for the target population of interest. At the IL, the Diabetes Prevention Program (DPP) is a Centers for Disease Control and Prevention recognized program delivered across the United States. The majority of the DPP encourages health behaviors that have been shown to be effective in not only preventing Type II diabetes mellitus, but also a number of other chronic disease conditions where overweight and psychosocial stress are contributing factors in their development or exacerbation. It has been adapted for high-risk groups with condensed sessions [41] and in community settings [42] with retained efficacy. Given this, the DPP was adapted to be disease-neutral, with other major format and goal changes outlined in Table 1.

**Table 1.**
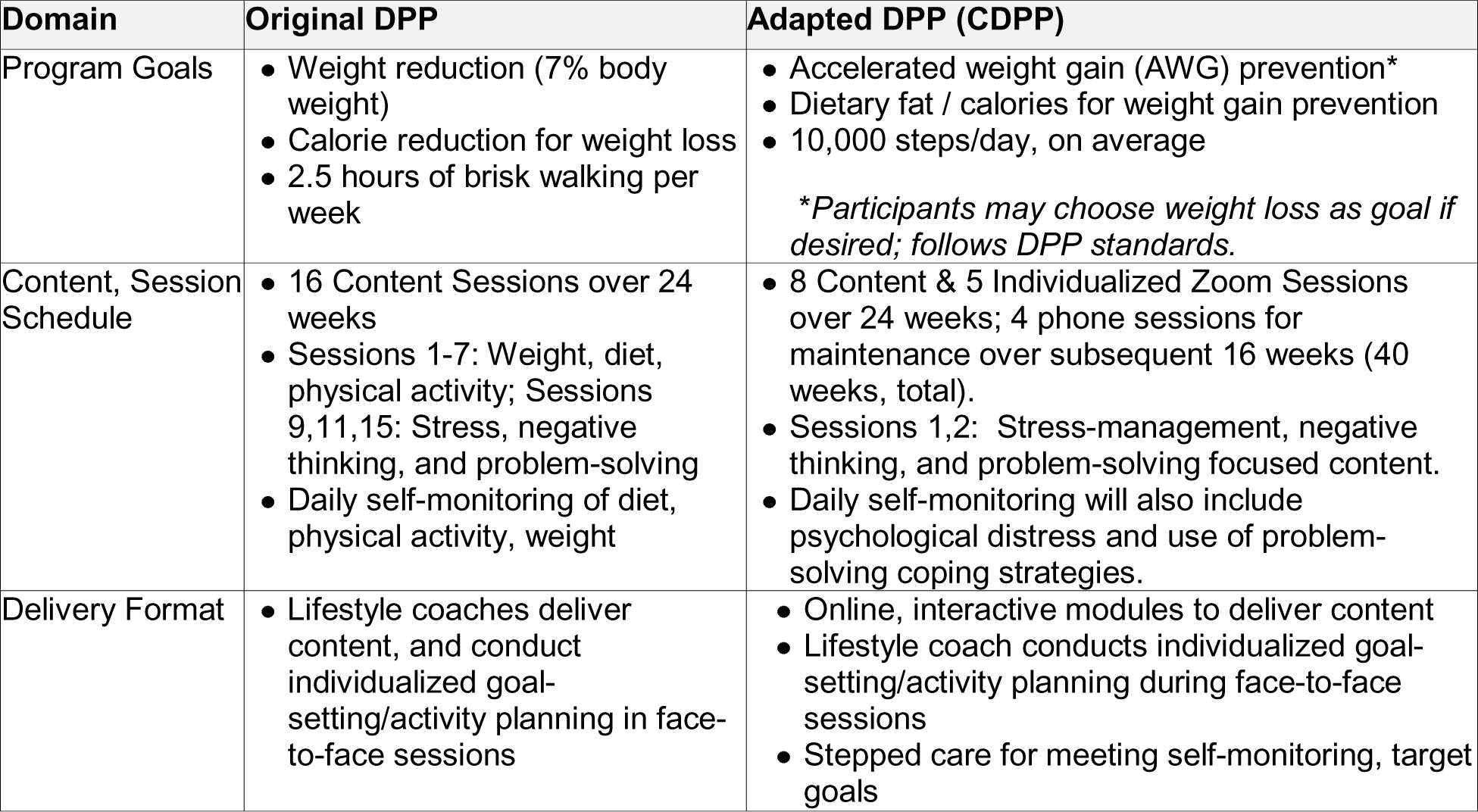
Diabetes Prevention Program (DPP) Adaptations.

| Domain | Original DPP | Adapted DPP (CDPP) |
| --- | --- | --- |
| Program Goals | • Weight reduction (7% body weight)<br>• Calorie reduction for weight loss<br>• 2.5 hours of brisk walking per week | • Accelerated weight gain (AWG) prevention*<br>• Dietary fat / calories for weight gain prevention<br>• 10,000 steps/day, on average<br>*\*Participants may choose weight loss as goal if desired; follows DPP standards.* |
| Content, Session Schedule | • 16 Content Sessions over 24 weeks<br>• Sessions 1-7: Weight, diet, physical activity; Sessions 9,11,15: Stress, negative thinking, and problem-solving<br>• Daily self-monitoring of diet, physical activity, weight | • 8 Content & 5 Individualized Zoom Sessions over 24 weeks; 4 phone sessions for maintenance over subsequent 16 weeks (40 weeks, total).<br>• Sessions 1,2: Stress-management, negative thinking, and problem-solving focused content.<br>• Daily self-monitoring will also include psychological distress and use of problem-solving coping strategies. |
| Delivery Format | • Lifestyle coaches deliver content, and conduct individualized goal-setting/activity planning in face-to-face sessions | • Online, interactive modules to deliver content<br>• Lifestyle coach conducts individualized goal-setting/activity planning during face-to-face sessions<br>• Stepped care for meeting self-monitoring, target goals |

Development of the CDPP occurred in partnership with community-based representatives who serve clients receiving DSS-E services (n=3), health department leaders (n=1), and clients receiving DSS-E services (n=3). Serving as an advisory board to the study team, this group met during two focus groups. The first focus group concentrated on general format (online, in person) options, order of content, length of learning modules and individualized attention from a lifestyle coach, and general approach for developing interactive components. A mock-up based on recommendations from the first focus group was generated by the Connected Health for Applications & Interventions (CHAI) Core of The North Carolina Translational and Clinical Sciences Institute (NC TraCS). The second focus group presented key features based on the group’s prior recommendations, eliciting details for developing 5 demographically diverse SED unemployed individuals as characters, each with their own life circumstances, stressors related to being unemployed and finding work, and health goals. The final intervention products and protocols were reviewed, commented on, and approved by prior focus group members for moving forward with adoption for use in the trial. Only minor adjustments were made following this last review and were focused on simplifying the intervention website to ensure it was user-friendly and expedited monitoring processes for end-users.

A more detailed crosswalk of how the CDDP maintained, and scheduled content from the original DPP as well as a popular adapted, condensed-session version (Group Lifestyle Balance, or GLB) [43] can be found in Table 2

**Table 2.**
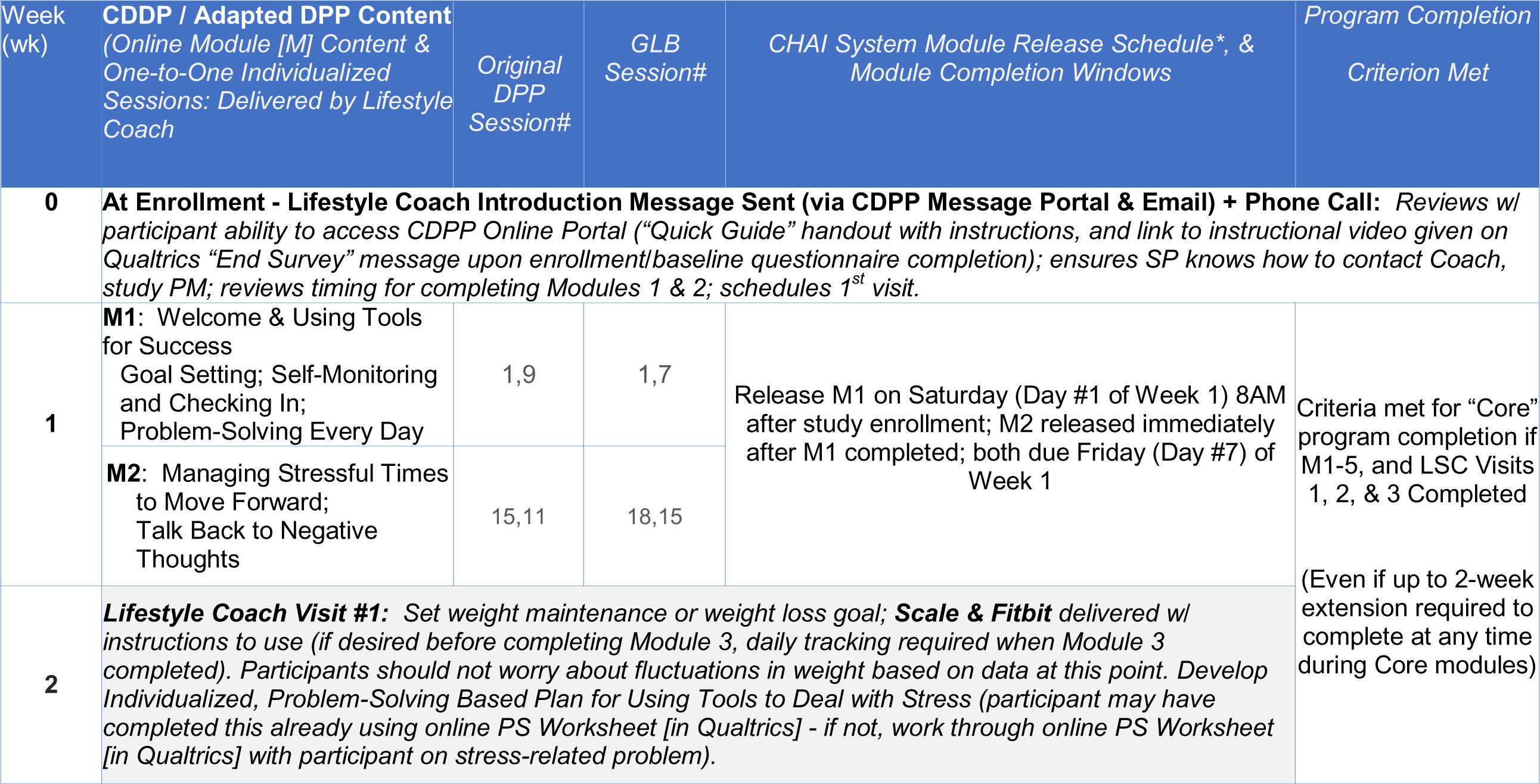
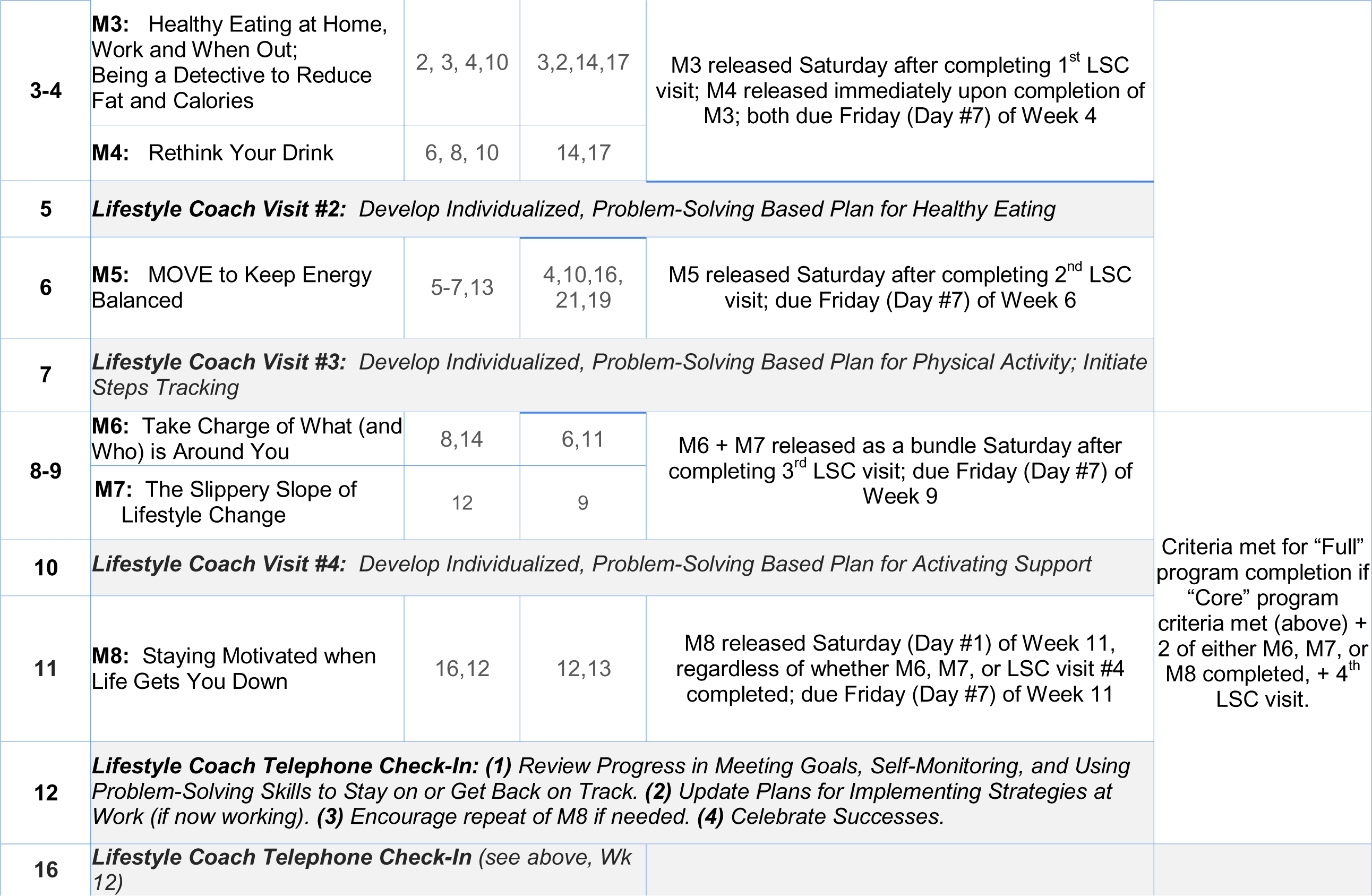
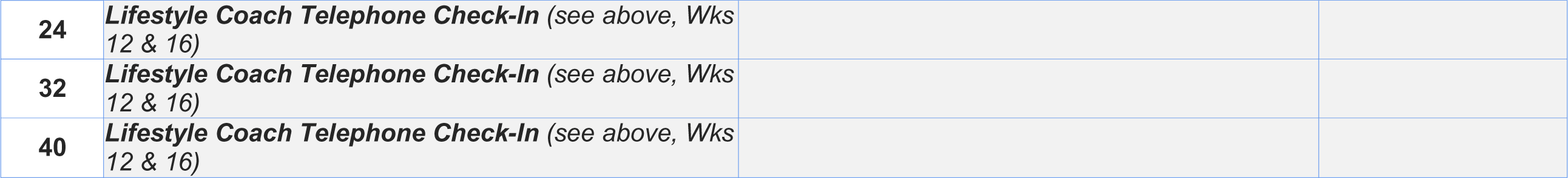
*NC Works4Health* CDPP Protocol, by Module & Lifestyle Coach Session, with Crosswalk to Original Diabetes Prevention Program (DPP) Content & Group Lifestyle Balance (GLB) Content.

#### Employer-Level Intervention: Supervising 4 Success (S4S)

The purpose of the intervention at the employer level was to mitigate the psychosocial stress newly-hired individuals can experience given their SED status and the many resource-deprivation challenges that status brings (e.g., reliable transportation, child care, etc.). As noted in Figure 1, an increase in psychosocial stress initiates a cascade of coping and health behaviors that can accelerate weight gain and increase other chronic disease risk. Using the categories described by Weiner and colleagues [44], the interaction of interventions at the IL and EL level are best described as a facilitation strategy, where continuing health-promoting lifestyle change at the IL after moving into a new work setting is facilitated by greater degrees of support in the work setting; in this case, from direct supervisors.

The S4S intervention was adapted from an implicit bias habit-breaking intervention developed by social psychologists and study collaborators at the University of Wisconsin-Madison [45, 46]. An advisory group of employer representatives / supervisors (n=10), and DSS-E representatives (n=2) who provide services for SED, unemployed adults was convened for two focus groups. The first focus group explored the acceptability of professional development focused on implicit bias and SED new hires; understandings of and preferences for terms used to describe implicit bias; format of delivery (online, in person); identifying real-life situations that should inform embedded exemplars; and frequency and timing recommendations for the training as well as the scheduled, 5-minute post-training “check-ins” between supervisors and new hires every week for 8 weeks, then every other week for the remaining 2 weeks to practice skills learned. A guide for topics to cover and questions supervisors can ask new hires to get to know them as individuals and provide needed support is provided as a link in the fourth module. A mock-up of a 5-module, online, 2.5 hour S4S training was reviewed with advisory group participants in a second focus group; minor adjustments were recommended to enhance interactive components and were integrated into the final intervention product for use in the trial. A certificate of completion is provided to individuals at the end of module completion.

**Figure 3.**
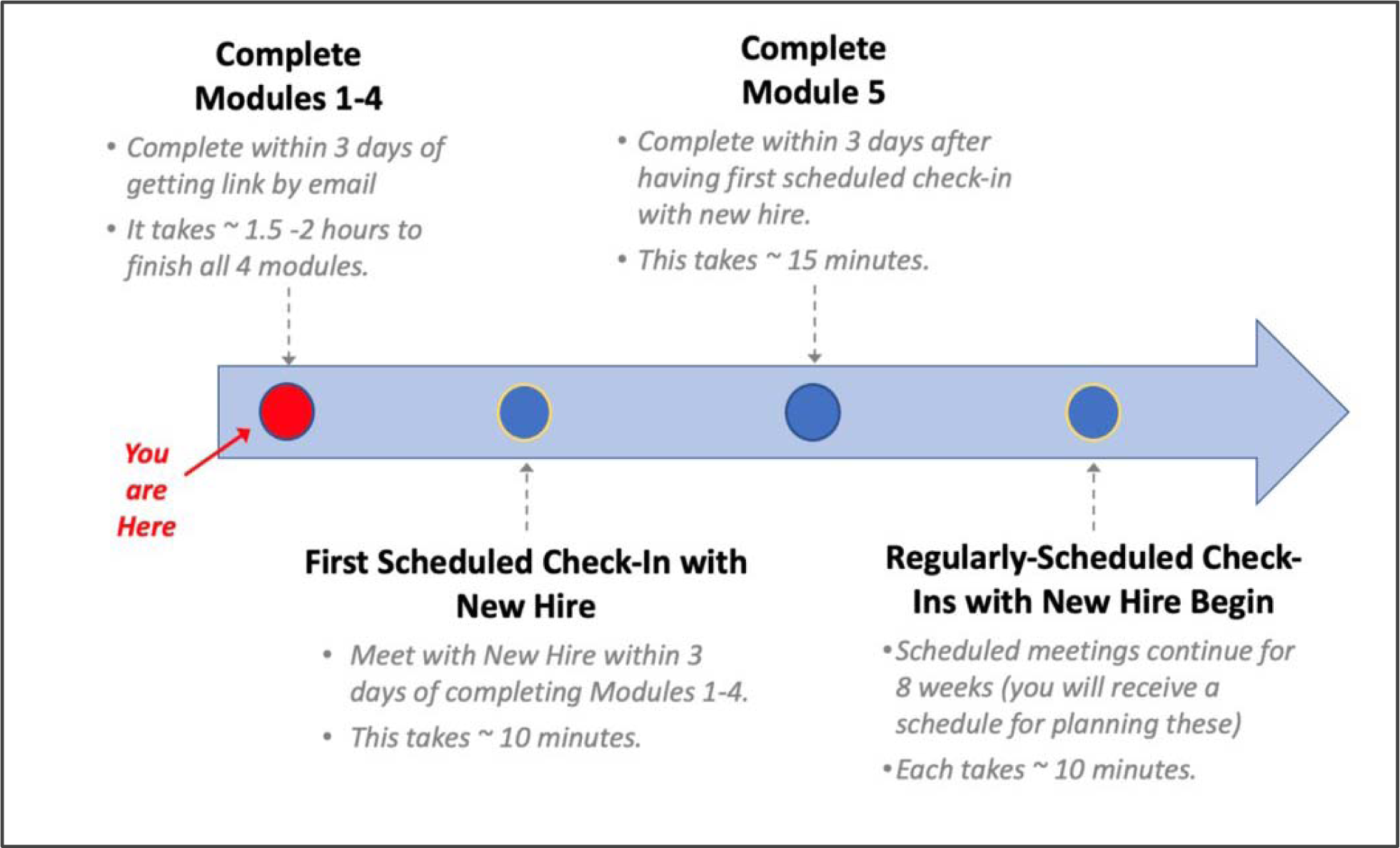
Adapted Supervising 4 Success (S4S) Intervention Program Schedule.

### Criteria for discontinuing or modifying allocated interventions {11b}

No harmful effects are anticipated; so there is no anticipated reason to stop the trial. In the event that there are clinically significant differences suggesting potential worsening of primary outcomes among the intervention group at the preliminary analysis point (when 300 IL participants have been enrolled), the study team will meet to review the findings and discuss whether to stop or continue the trial. A change in allocation would only occur at the employer-level, such that there was significant imbalance in supervisors enrolled from large (500 or more employees) or small (less than 500 employees) employers.

### Strategies to improve adherence to interventions {11c}

The intervention adaptations were designed with representatives of the study target populations and other relevant stakeholders during focus groups. Final intervention materials were reviewed by these groups with all participants deeming the intervention acceptable for use in the trial. Intervention formats were selected to enhance easy access independent of study participant schedules or transportation access.

### Relevant concomitant care permitted or prohibited during the trial {11d}

There were no restrictions of concomitant care at either the individual or employer levels. Given the frequent scarcity of resources among SED unemployed individuals, study participants were given a list of community resources for meeting basic housing, food, safety, and health-related needs. Although many unemployed study participants were receiving individual case-management services through public agencies, with the employer’s permission, we also provided contact information for employers enrolled in the trial who had open positions.

### Provisions for post-trial care {30}

No provisions were made for post-trial care, as there is no anticipated harm from participation.

### Outcomes {12}

Primary study outcomes include change over time in weight gain, blood pressure, and psychological distress. Subject-specific linear versus quadratic growth models for each outcome using generalized multilevel models with an appropriate link function to account for response distributions of the outcomes will be compared for main effects at the individual and employer level, as well as the interaction effects. Clustering of participants at the IL hired within employers will be handled with a sandwich estimator to generate robust standard errors. Secondary outcomes include situational stress, coping style, health self-regulation, dietary intake, physical activity, perceived supervisor support, work limitations, and employment duration.

### Participant timeline {13}

Participant recruitment for the trial began in September 2021 and is anticipated to be complete by December 31, 2024. Participants at the individual level remain in the trial for 12 months, with data collection at baseline, 3 months, 6 months, and 12 months; intervention participants have more intensive study involvement for 40 weeks after enrollment with the CDPP intervention delivery. Employers remain in the trial until all data collection is complete.

### Sample size {14}

We will enroll 600 unemployed SED adults at the individual level, assuming a minimum of 420 of these individuals will become employed with one of the enrolled, randomized employers. At the EL, any supervisor assigned to supervise one of the IL study participants will be recruited into the study, anticipating enrolling up to 400 supervisors (including potential supervisor turnover during the study period) and up to 200 employers. We conducted a Monte Carlo simulation study using 100 replications generated using MLPowSim[47] to determine the expected power on each primary outcome as continuous measures (psychological distress, weight gain, blood pressure) given an employer-level ICC of .05. Based upon prior literature [48–51], we assumed a Cohen’s *d* of .3 for both of the main effects in each primary outcome. The individual-level intervention (alone) or the employer-level intervention (alone) would have a Cohen’s *d* of .3, but the combined interventions would have a Cohen’s *d* of .6. Under these conditions, we will have power of .975 to detect the main effects of the individual-only and employer-only intervention groups and >99% power to detect the combined intervention effect.

### Recruitment {15}

IL study participants are recruited using a variety of community-wide approaches, including social media platforms; study advertisements posted in local job-search agencies; at job fairs and other employment-oriented community events; through email listservs held by DSS-E partnering agencies; and through other community agencies serving unemployed, SED populations (churches, food pantries, etc.). Employers are recruited through many of the same means as IL participants; in addition, we have engaged local Chambers of Commerce in study counties to assist recruiting employers.

At the start of the trial, supervisor recruitment was initiated after identifying the supervisor and the employing organization they worked for, enrolling the employer organization when the IL study participant found employment, and then approaching the supervisor to enroll. Post-COVID labor market changes made this approach exceedingly difficult, and recruitment for supervisors and employers was modified in early 2023 after convening local employers to assist with developing new employer and supervisor recruitment and enrollment strategies. Recommendations from this local employer advisory group were to 1) offer all employers and their supervisory staff free access to the S4S intervention in a shorter time frame (rather than randomizing them to a control condition without the ability to access S4S until study completion several years later), given the dire, post-COVID-19 need to retain workers; 2) emphasize the diversity, equity, and inclusion (DEI) content embedded in S4S, given many employers are spending thousands of dollars for DEI training compiled by private vendors; and 3) emphasize the evidence-based nature S4S, given trainings by private vendors may or may not be evidence- or theory-based.

Based on these recommendations, changes to employer and supervisor recruitment and enrollment were made in March, 2023. We are now enrolling as many employer organizations as possible in study counties, and prioritizing recruitment efforts on employers with existing relationships and that hire job seekers from DSS-E programs – increasing the likelihood our IL study participants would be hired from these organizations. Each employer organization is randomized using a stepped wedge approach to either receive the intervention for all supervisors immediately, in 3 months, or in 6 months, without being dependent on an IL study participant being hired into the organization first. As such, our IL study participants hired by enrolled employers will have supervisors that would already be randomized to the wait-control period (if assigned to to receive access to the S4S intervention in 3 or 6 months). Supervisors of IL study participants who are not already embedded in an enrolled employer organization are randomized to either receive the S4S intervention or to the control group as individuals, randomizing to group in blocks of four.

## Assignment of interventions: allocation

### Sequence generation {16a}

Randomization sequences are computer-generated at all levels (IL SED unemployed adults; employer organizations; and individual supervisors when not embedded within enrolled employer organization) using a maximal imbalance method.[52, 53] This approach enforces strict balancing similar to permuted block randomization but samples from random walks that satisfy the imbalance restrictions rather than blocks which make the allocation sequence less predictable. The randomization sequences were generated in R version 4.1.1 using the MPBoost package [54], and the randomization seed was set to the most recent North Carolina Pick 4 Lottery drawing at the time the sequence was generated, which was the evening draw on September 26th 2021.

### Concealment mechanism {16b}

Allocation to group is revealed only after each unit is enrolled (individuals, employer organizations, or supervisors). The randomization sequence is inaccessible to all members of the study team and cannot be viewed prospectively, as the sequence is generated remotely and allocation to group occurs only in real time, at the moment of each unit’s enrollment.

### Implementation {16c}

Study participants enrolling at the IL complete informed consent and the baseline questionnaire using Qualtrics survey software. A random assignment generator is embedded in the final section of the survey; study participants are informed of their group allocation (i.e., the survey notification language refers to the “study control group” or the “Chronic Disease Prevention Program group”). Qualtrics surveys are completed by the study recruitment coordinator to randomize employer organizations and supervisors immediately after they enroll, with each survey having embedded random assignment generators as noted previously. Study staff are notified of the group assignment from within the Qualtrics survey, and inform the newly-enrolled employment organization representative or the supervisor, as appropriate.

## Assignment of interventions: blinding

### Who will be blinded {17a}

Once participants are recruited and randomized, masking their allocation is not possible. Data collectors are not notified of participants’ group assignment; however, study participants often reveal their group assignment to them when they meet. The study biostatistician is blinded to group when conducting preliminary outcome analyses as needed for periodic Data, Safety, and Monitoring Committee (DSMC) review.

### Procedure for unblinding if needed {17b}

Preliminary analyses of main and interaction effects for primary outcomes that are potentially clinically significant will elicit unblinding of group assignment; should the data suggest potentially negative effects of the interventions at either level, the research team will meet to discuss the findings and make decisions around continuing the trial or making other modifications to trial protocols to ensure safety of the trial participants. All findings and plans will be reported to the university Institutional Review Board.

## Data collection and management

### Plans for assessment and collection of outcomes {18a}

All study outcomes are measured at the IL. Primary study outcomes include psychological distress, weight, and blood pressure; secondary outcomes include situational stress, coping style, health behaviors, perceived workplace support, health-related employment functioning, and employment duration. Measures used for each outcome are detailed in Table 3.

**Table 3.**
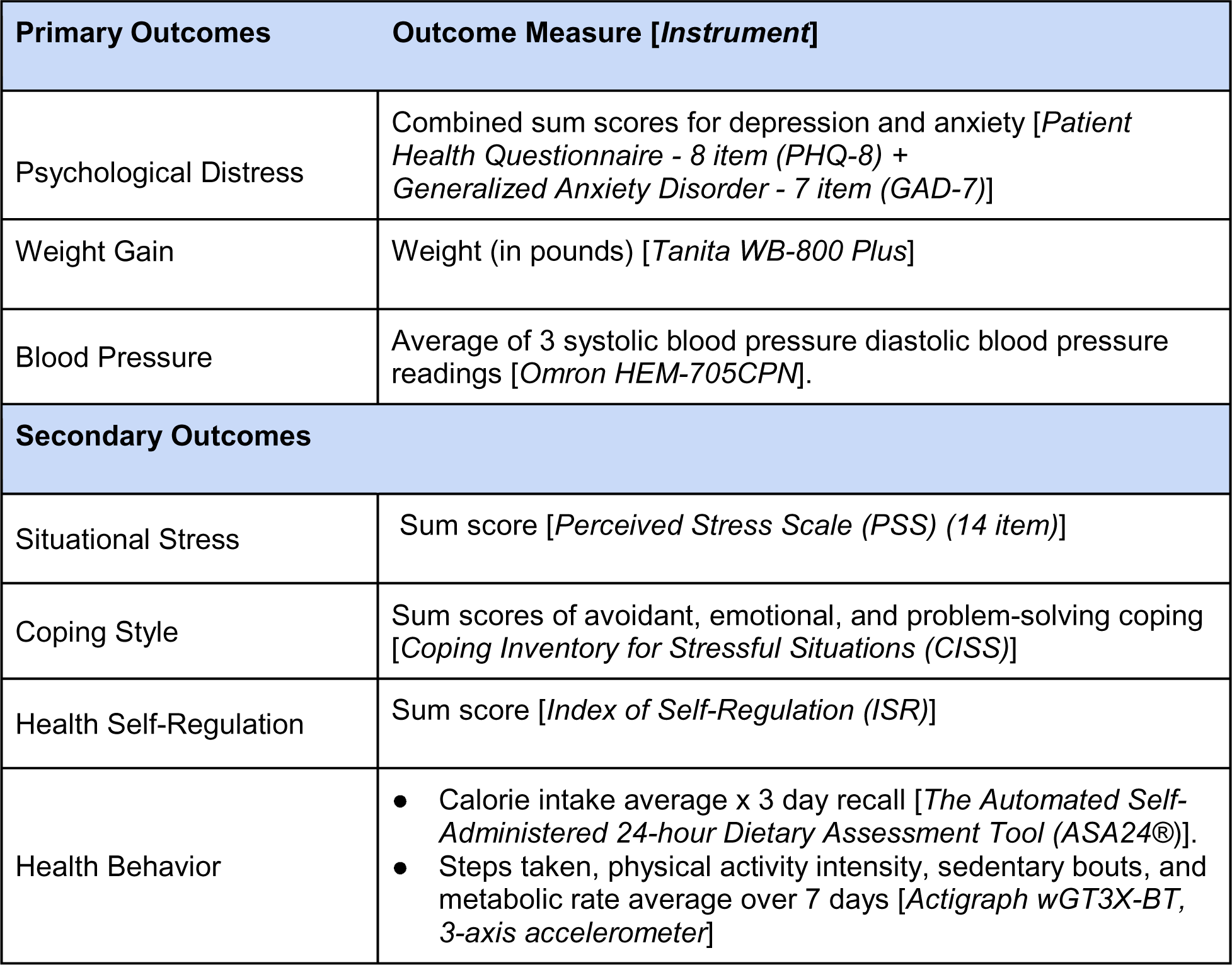
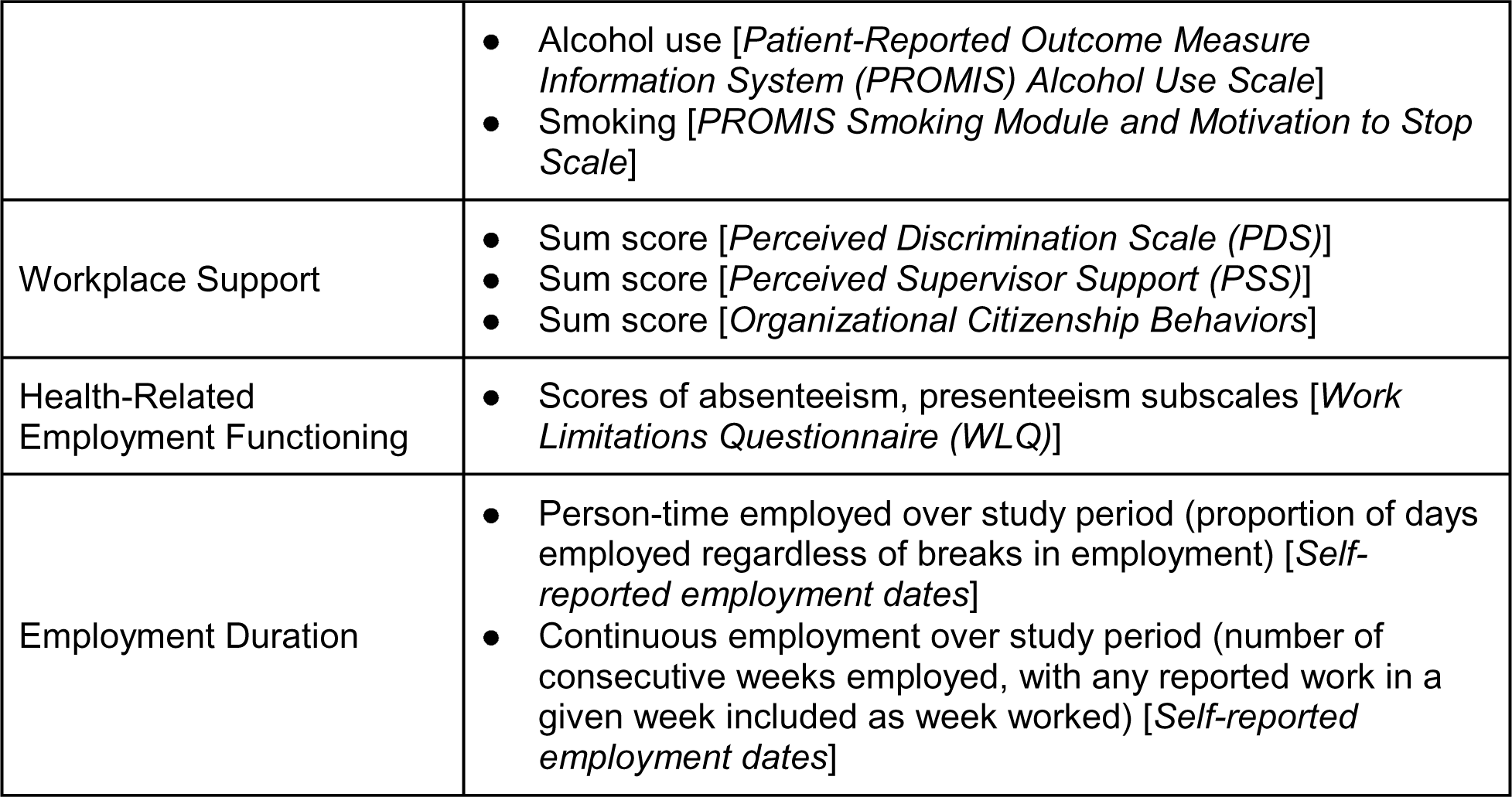
Primary and Secondary Outcome Measures.

| Primary Outcomes |  | Outcome Measure [*Instrument*] |
| --- | --- | --- |
| Psychological Distress |  | Combined sum scores for depression and anxiety [*Patient Health Questionnaire - 8 item (PHQ-8) + Generalized Anxiety Disorder - 7 item (GAD-7)*] |
| Weight Gain |  | Weight (in pounds) [*Tanita WB-800 Plus*] |
| Blood Pressure |  | Average of 3 systolic blood pressure diastolic blood pressure readings [*Omron HEM-705CPN*]. |
| Secondary Outcomes |  |  |
| Situational Stress |  | Sum score [*Perceived Stress Scale (PSS) (14 item)*] |
| Coping Style |  | Sum scores of avoidant, emotional, and problem-solving coping [*Coping Inventory for Stressful Situations (CISS)*] |
| Health Self-Regulation |  | Sum score [*Index of Self-Regulation (ISR)*] |
| Health Behavior |  | • Calorie intake average x 3 day recall [*The Automated Self-Administered 24-hour Dietary Assessment Tool (ASA24®)*].<br>• Steps taken, physical activity intensity, sedentary bouts, and metabolic rate average over 7 days [*Actigraph wGT3X-BT, 3-axis accelerometer*] |

|  |  |
| --- | --- |
|  | Alcohol use [*Patient-Reported Outcome Measure Information System (PROMIS) Alcohol Use Scale*]<br>Smoking [*PROMIS Smoking Module and Motivation to Stop Scale*] |
| Workplace Support | Sum score [*Perceived Discrimination Scale (PDS)*]<br>Sum score [*Perceived Supervisor Support (PSS)*]<br>Sum score [*Organizational Citizenship Behaviors*] |
| Health-Related Employment Functioning | Scores of absenteeism, presenteeism subscales [*Work Limitations Questionnaire (WLQ)*] |
| Employment Duration | Person-time employed over study period (proportion of days employed regardless of breaks in employment) [*Self-reported employment dates*]<br>Continuous employment over study period (number of consecutive weeks employed, with any reported work in a given week included as week worked) [*Self-reported employment dates*] |

At the IL, data collection occurs at baseline, and again at 3-, 6-, and 12-months after enrollment. All self-report data are collected using an online survey developed using Qualtrics Survey Software [55]. Qualtrics was also used to develop a data collection protocol that serves both as a data collection tool (data collectors enter weight and blood pressure readings into the survey during the visit) and a tool to facilitate and monitor fidelity to the in-person data collection protocol. All data collectors were trained and monitored regularly by a study staff member responsible for all data collection activities. At the in-person data collection visit, data collectors assist study participants in completing the online self-report questionnaire if they have not yet done so, demonstrate use of the ASA24^®^ online reporting process, and remind participants of how to wear and care for the actigraph, which participants were instructed to wear on their wrist and if unable, were fitted for and instructed in wearing around their waist.

### Plans to promote participant retention and complete follow-up {18b}

Select aspects of the study were designed to promote retention and follow-up. In the formative research conducted prior to the trial, stakeholder groups representing the target populations (SED, unemployed adults, supervisors, and employers) were key collaborators in guiding the intervention adaptations at each level to facilitate intervention acceptability, uptake, engagement, and completion.

At the IL, to maximize in-person trust-building with study personnel, data collectors were assigned to IL participants at the time of enrollment and completed all follow-up data collection, unless there was a specific need to reassign data collectors. Other consistent points of contact for the study include the study manager and recruitment coordinator – each of whom have designated groups to contact to facilitate continuity and study recognition. Gift card incentive amounts were increased at each data collection point ($35 at baseline; and $40, $50, and $60 for completing data collection at the 3-, 6-, and 12-month follow-up occasions, respectively). Finally, IL participants allocated to the control group are given the option of receiving access to the CDPP modules and online health behavior tracking resources for use after completing their 12-month data collection.

At the EL, there was also consistency in terms of study personnel for employers to contact. Given the stepped-wedge randomization could result in a 3- or 6-month delay in accessing the S4S intervention, monthly automated email messages were sent to designated contacts within employer organizations reminding them of when they will have access to the intervention materials for their supervisors.

### Data management {19}

Secure software platforms that allow for shared use of tracking documents and access to study data facilitate dataset and data collection version control processes (Qualtrics Survey Software [55], Microsoft Teams [56], and a UNC-CH internally-controlled intervention delivery website). Study identification numbers (IDs) are assigned using an automated random number generator within Qualtrics when participants begin the study registration and enrollment process, stored, and pulled into all other data collection surveys when the participant enters their email address for authentication. Study IDs are reviewed on a regular basis to assess for duplicate ID assignments and make corrections, where necessary. Stata/SE Version 17.0 is used to clean and analyze the data using the “.do” file feature to ensure cleaning, management, and other analytic commands are applied consistently [57].

### Confidentiality {27}

Confidentiality for participants at both levels comply with Institutional Review Board standards at The University of North Carolina at Chapel Hill.

### Plans for collection, laboratory evaluation, and storage of biological specimens for genetic or molecular analysis in this trial/future use {33}

No biological or genetic specimens will be collected in the study.

## Statistical methods

### Statistical methods for primary and secondary outcomes {20a}

**<u>Aim 1</u>** will assess the main effects of the employer-level and individual-level interventions on primary outcomes (psychological distress, weight gain, and blood pressure) and secondary outcomes over time. **<u>Aim 2</u>** will assess the interactive effects of the interventions across levels. Our primary analysis will be conducted with DSS-E hires nested within these employers. After performing exploratory data analysis including data visualization and descriptive statistics,we will fit separate marginal models for each of the three primary outcomes with baseline measurements, a linear calendar time trend, and linear time on intervention (including the time on both individual- and employer-level interventions interaction) as predictors. The models will be estimated by generalized estimating equations [58] with a block exchangeable correlation structure and robust standard errors. The correlation structure will be estimated using matrix-adjusted estimating equations [59] because of their potential to improve finite-sample performance and greater robustness to small cluster sizes at the employer level. We will conduct sensitivity analyses with categorical and higher-order polynomial parameterizations of the calendar time trend and the time on the interventions.

We will estimate the marginal participant-average treatment effects in terms of the risk differences for the primary outcomes after 12 months of treatment. Confidence intervals will be computed and adjusted to control the false discovery rate using the Benjamini-Yekutieli procedure [60] to account for multiple comparisons i.e. the three models and, for the primary analysis, the two separate intervention parameters of interest for the individual- and employer-level interventions respectively. While an interaction term is included in the model, no hypothesis tests are conducted with it for the primary analysis. A success on any outcome (adjusted for multiple comparisons) will be considered a success for the intervention; that is the interventions do not need to significantly improve all outcomes to be considered a success.

### Methods for additional analyses (e.g., subgroup analyses) {20b}

We will use an intent-to-treat approach for our primary analyses. Subgroup analyses of main intervention effects by CDPP intervention completers, S4S intervention module completers, level of supervisor support, and gender will be conducted for exploratory purposes only.

Missing outcome data will be handled using full information maximum likelihood (FIML) estimation under the assumption that missing data are missing at random (MAR) conditional on observed outcome data and on observed predictors. The benefit of this approach is that it can handle erratic, wave-level missingness as well as dropout. Missing data on predictors will be handled using multiple imputation. Multilevel multiple imputation will be used to impute employee-level predictors and standard multiple imputation will be used to impute employer-level predictors. Finally, we will apply a Complier Average Causal Effect (CACE) analysis if > 5% contamination occurs to address this as a source of potential bias [61–64].

### Methods in analysis to handle protocol non-adherence and any statistical methods to handle missing data {20c}

Routine observation of the CDPP intervention delivery occurs every 3 months. The S4S intervention module completion can be determined using Qualtrics online survey data capture. Substantive protocol non-adherence is reported to the university IRB, remedial training for and more frequent monitoring of study team members is scheduled, and modifications or clarifications to the protocols are made as needed. Missing data will be accounted for using multilevel multiple imputation with chained equations [65].

### Plans to give access to the full protocol, participant-level data, and statistical code {31c}

At the end of the study, the team will prepare a final, de-identified limited-access dataset suitable for sharing with other researchers. De-identification will include removal of obvious identifiers such as names and addresses. It will also include examination of less obvious potentially identifying variables. Continuous variables with extreme values may have the extremes truncated. Height and weight are examples of such variables. Categorical variables with small numbers of participants in some categories may have these categories pooled with larger categories.

The limited-access dataset will be made available at the end of the study. Data will be released in a .csv file. Researchers wishing to use the dataset will be required to obtain approval from UNC-CH using an agreement form that clearly specifies the intended analyses as well as assurances that HIPAA regulations will be followed. If approval is granted, the dataset will be supplied on a CD, or shared using a secure, password-protected medium. It will also include documentation describing the variables in the dataset, Stata/SE Version 17.0 command codes for cleaning and analyses, and copies of the data collection forms used to collect the data [57].

## Oversight and monitoring

### Composition of the data monitoring committee, its role, and reporting structure {21a}

Given the low risk of study participation in both the individual level and employer level intervention arms, a Data Safety and Monitoring Board is not needed; however, a Data Safety and Monitoring Committee (DSMC) to provide additional data management oversight includes the study statistician, data manager, and select members of the academic research team.

### Adverse event reporting and harms {22}

All adverse events are reported to the IRB. Potential harms include unintended disclosure of identifying information and financial repercussions from the employers/supervisors; all other potential harms are anticipated to be minor.

### Dissemination plans {31a}

Dissemination of results will include paper and poster presentations at scientific nursing and public health conferences; publications related to study processes and outcomes. Recruitment and retention outcomes are regularly communicated to our partner organizations, and findings at the end of the study period will be shared with community partners in in-person forums and through written communication, regardless of whether the outcome findings are negative, positive, or null. Interventions were developed with implementation in mind; should findings be positive, dissemination will include effective strategies to enhance adoption of interventions at the local, state, and federal levels.

## Discussion

Researchers working in the field of public health understand the role that social determinants of health (SDOH) play in driving health inequities. There is a clear imperative to intervene on employment as one of several SDOH if we are to make any real progress in mitigating health inequities. Developing and testing interventions across multiple levels that include cross-sector partnerships is fundamental to these efforts [39, 66]. Efforts to advance designing MLIs that produce synergistic effects [44] are ongoing and fraught with methodological challenges [66]. As noted in the *NC Works 4 Health* study example, intervening on SDOH related to employment is nuanced and requires the engagement of many different actors across sectors, settings, and time. Moreover, as higher rates of precarious work for SED populations increase in the U.S. labor market [67], conditions remain ripe to widen health inequities without intervention. Finally, trials testing MLIs that intervene at critical points along the natural trajectory of SED populations transitioning into and out of employment, and across employment settings, are needed. Such trials will require novel approaches for engaging employers and supervisors in the work, which has historically been challenging, at best [68].

## Trial status

The trial is in the recruitment and intervention delivery phase, with 248 IL study participants and 16 employers enrolled to date.

## Abbreviations

SED: Socioeconomically disadvantaged
DSS-E: Department of social services employment
IL: Individual level
EL: Employer level
UNC-CH: University of North Carolina at Chapel Hill
ID: Study ID
HIPAA: Health Information Protection and Accountability Act
DSMC: Data Safety and Monitoring Committee
CDPP: Chronic Disease Prevention Program
DPP: Diabetes Prevention Program
S4S: Supervising for Success
DEI: Diversity, Equity, and Inclusion
NCWorks4Health: North Carolina Works for Health Study
IRB: Institutional Review Board

## Ethics Approval

The NCWorks4Health study was approved by the University of North Carolina at Chapel Hill Non-Biomedical IRB (Ref: 21-0859). The study is being conducted in accordance with the ethical standards as laid down in the 1964 Declaration of Helsinki and its later amendments.

## Consent to Participate

Informed consent was obtained from all individual participants included in the study.

## Data Availability

All data produced in the present study will be available upon reasonable request to the authors within 4 months of study completion.

